# Plasma proteins are integral to gene-regulatory networks acting within and across blood cells, the arterial wall and major metabolic organs

**DOI:** 10.1101/2025.01.22.25320723

**Authors:** Sean Bankier, Husain Talukdar, Mariyam Khan, Giuseppe Mocci, Katyayani Sukhavasi, Ke Hao, Angela Ma, Arno Ruusalepp, Eric E Schadt, Jason C. Kovacic, Tom Michoel, Johan LM Björkegren

## Abstract

The plasma proteome is the future for diagnostic markers for common diseases, like cardiometabolic disorders (CMDs) and coronary artery disease (CAD). The reliability of these markers requires identifying their source-organ(s). We profiled 974 plasma proteins in 532 CAD-patients of the STARNET study with arterial wall, major metabolic organ, and blood transcriptomic data. 144 plasma cis-pQTLs colocalized with tissue eQTLs including eight CMD/CAD GWAS genes. 262 plasma proteins correlated with their corresponding tissue “seed” genes whereof 101 in the liver. 851 plasma proteins were strongly associated with the activity of gene-regulatory networks (GRNs), particularly cross-tissue GRNs. The Adipose-Liver Plasma LeptIN-regulating GRN78 with the top key driver *UCHL1* in fat stood out. Beyond genetics, explaining up to 20% of plasma protein variation, and a limited number of mostly hepatic seed genes, plasma proteins are integral to GRNs acting within and across blood cells, the arterial wall and major metabolic organs.

## INTRODUCTION

The human plasma proteome is defined by the total number of proteins that at a given time point are found in circulating blood. These “plasma” proteins can arise from various tissues throughout the body following their secretion, or leakage from damaged cells^1^. Moreover, unlike proteomes within cells of a given organ system (e.g., the liver) that largely define their physiological functions, plasma proteins do not represent a coherent function but rather reflect the overall status of organs throughout the body^2,3^. To what extent the functions of given organs are reflected by specific plasma proteins or whether the levels of plasma proteins jointly reflect the functions of several organ systems remain largely unexplored. Up to 20% of the variation in plasma protein levels is explained by common genetic variation ^2,4^. Therefore, besides the potential of plasma proteins to reflect conditions of healthy organs, genome-wide association studies (GWAS) have, by colocalization analyses, linked genetic variation of plasma protein levels^2,4^ to the genetic risk of developing diseases including several common cardiometabolic disorders (CMDs) such as obesity^5^, coronary artery disease (CAD)^6,7^, type II diabetes and stroke^8^.

Given that most plasma proteins are secreted or leaked into the circulation following their synthesis in various organs and cells, the genetic regulation of plasma proteins (i.e., protein quantitative trait loci (pQTLs)) should largely reflect that of gene expression (e)QTLs within and across organs. Indeed, evidence of such colocalization between tissue-specific eQTLs in GTEx^9^ and plasma pQTLs in UK Biobank was recently demonstrated^2^. Besides genetic variation, other factors influencing the levels of plasma proteins include micro-environmental factors affecting transcription and translation across tissues prior to their secretion, post-translational modifications and plasma clearance. To better understand both the genetic and non-genetic regulation of plasma proteins, we measured the levels of 974 plasma proteins in 532 coronary artery disease (CAD) patients of the well-characterized, CMD-focused Stockholm-Tartu Reverse Networks Engineering Task (STARNET) study^10,11^. In addition to blood and plasma, STARNET uniquely contains parallel samples from the liver, skeletal muscle, subcutaneous and visceral abdominal fat and the atherosclerotic and non-atherosclerotic arterial wall from living CAD patients undergoing coronary by-pass surgery. We found that besides genetic variation in the form of eQTL/pQTL pairs and 161 plasma proteins linked to the expression of their coding genes (“seed genes”), gene regulatory networks (GRNs) acting within and across major metabolic organs and the arterial wall are major determinants of the levels of plasma proteins.

## RESULTS

### Genetic regulation of plasma proteins colocalizes with that of cardiometabolic gene expression and traits

Applying a cis-window of ±500Kb around the transcription start sites for a total of 945 genes corresponding to the measured plasma proteins, 1,860,901 SNPs were identified. Of these, 50,106 SNPs were identified as cis-pQTLs for 708 unique proteins in plasma (FDR=5%) (Supplementary Table 2). Next, using the same set of 1,860,901 SNPs, eQTLs for 828 unique genes were identified in at least one of the seven tissues, including the 708 genes corresponding to the plasma pQTLs (Figure 1). Per tissue, the corresponding gene numbers with eQTLs were 634 in visceral abdominal fat, 617 in subcutaneous fat, 610 in liver, 596 in the atherosclerotic arterial wall, 560 in the non-atherosclerotic mammary artery, 484 in whole blood and 440 in skeletal muscle. To examine the extent by which the regulatory effects of eQTLs for the “seed genes” which encode (Methods) plasma proteins are transmitted to also form pQTLs (Figure 2A), we examined the overlap between eQTLs across the seven tissues with that of pQTLs (Figure 2B). eQTLs acting across all seven or six tissues had the highest overlap with pQTLs, followed by tissue-specific eQTLs in blood, liver and the atherosclerotic arterial wall (AOR) (Figure 2B). Across tissues, the allelic effect direction for the seed genes and plasma protein was for the most part the same (80-90%, Figure 2C) exemplified by FOLR3 (blood) and MICA (SKLM). In contrast, IL6R (MAM) was a prominent example of opposite allelic effects (Figure 2D). For pQTL-overlapping eQTLs acting across several tissues, the beta values of the tissue-specific eQTLs were mostly coherent (Figure 2E).

**Figure 1.**
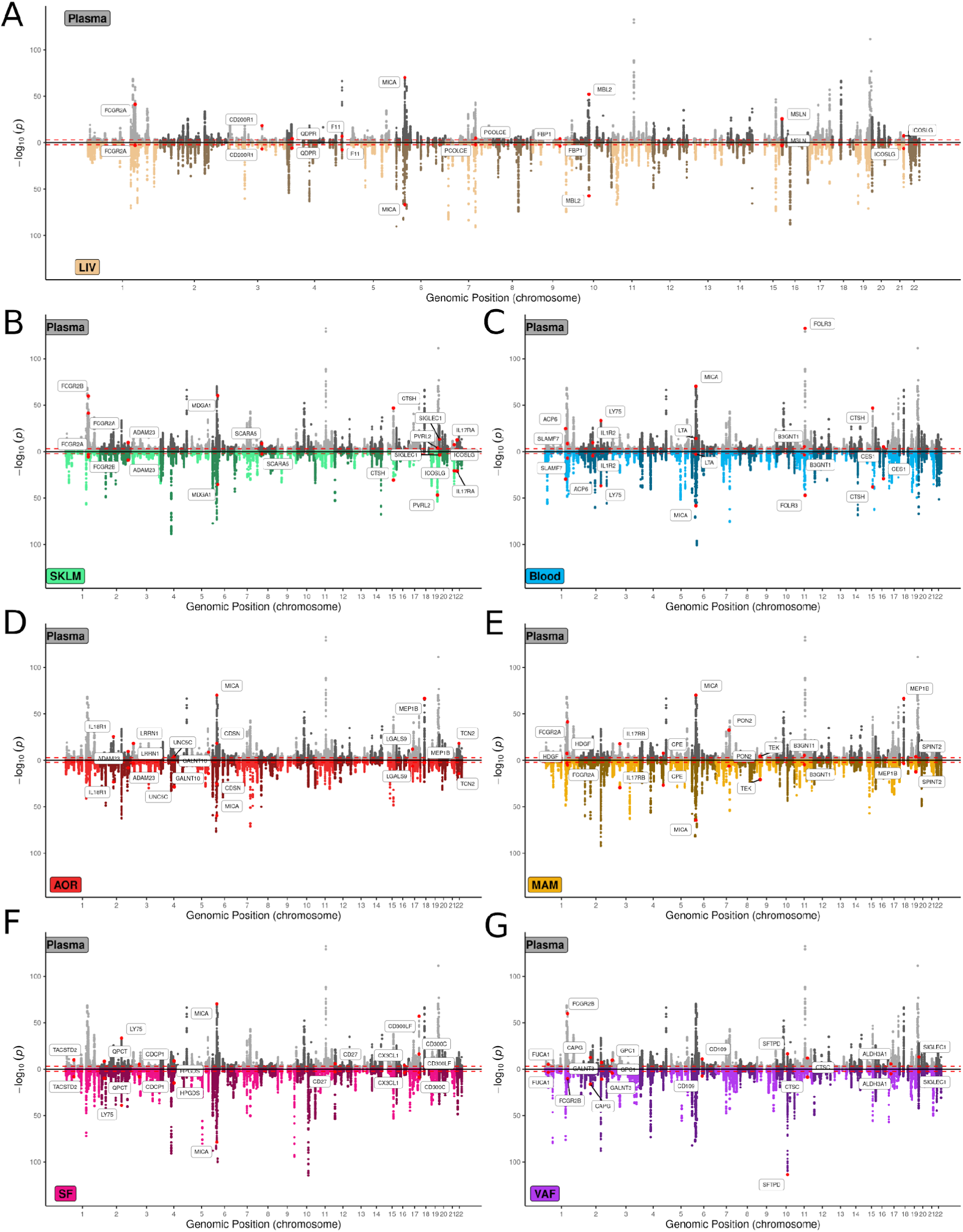
Manhattan plots of plasma cis-pQTLs mirrored by corresponding tissue-specific cis-eQTLs. **A-G**, Mirror Manhattan plots showing genomic location of cis-pQTLs (± 500Kb TSS) for 50,107 pSNPs (FDR=5%) of 945 proteins measured in plasma from 461 STARNET participants (upper panels) mirrored by corresponding tissue-specific cis-eQTLs (lower panels) in liver (A), skeletal muscle (B), blood (C), atherosclerotic aortic root (D), internal mammary artery (E), subcutaneous fat (F) and visceral abdominal fat (G)^10,11^. cis-pQTL/cis-eQTL pairs indicated with names are the top 10 most significant pairs for each tissue.

**Figure 2.**
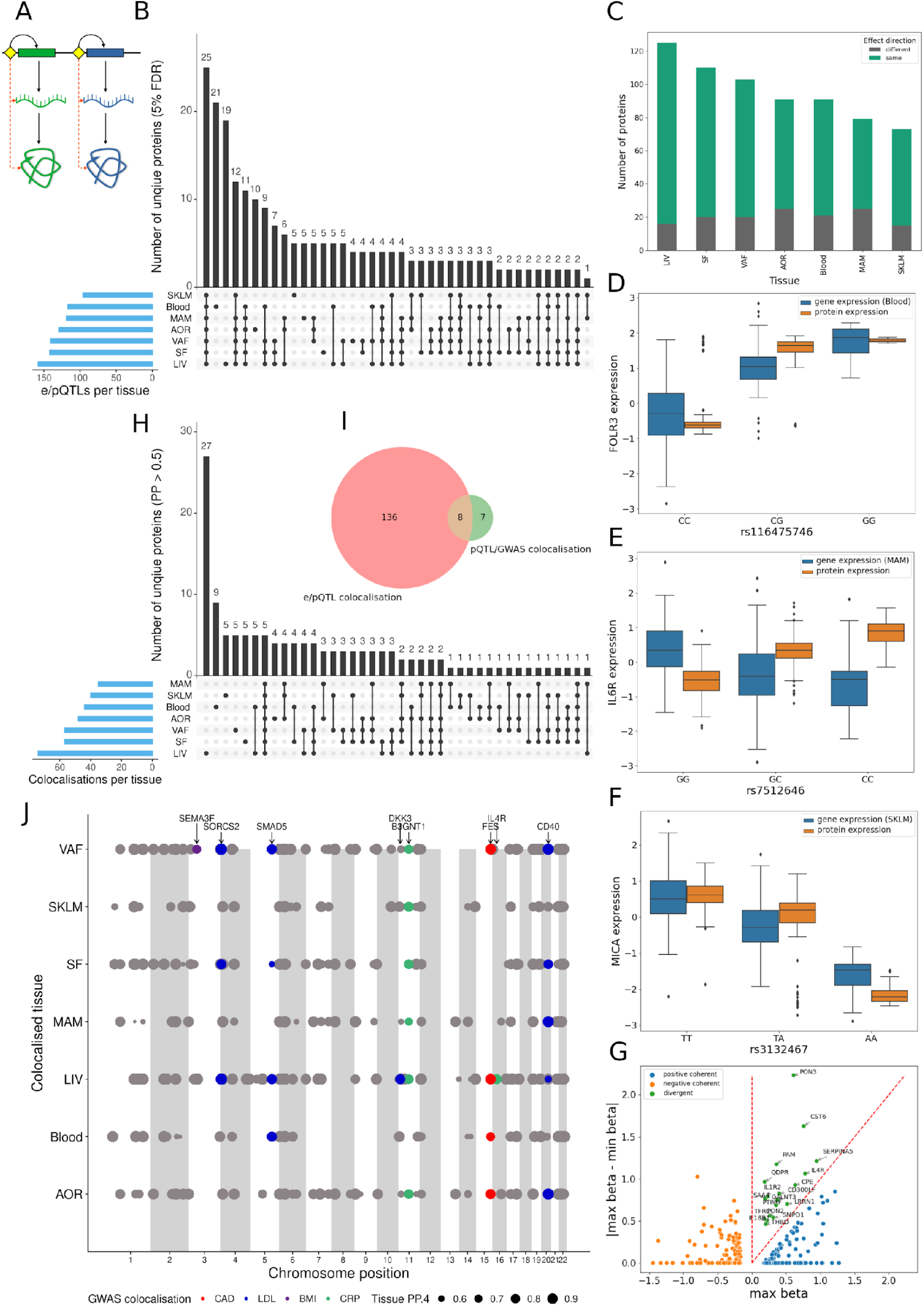
Overlap and co-localization of cis-eQTLs in metabolic and arterial wall tissues with corresponding plasma cis-pQTLs in relation to cardiometabolic risk loci identified by GWAS. **(A)** Schematic illustration showing the principal hypothesis that the genetic regulation of seed gene expression (i.e., mRNA) levels in tissues (i.e., cis-eQTLs) is transmitted to also regulate the levels of corresponding proteins in plasma (i.e., cis-pQTLs). Black arrows, flow of information; dashed red arrows, measured correlations. **(B)** Upset plot showing the numbers of plasma proteins that share genetic regulation with that of their seed genes across tissues (i.e., cis-eQTLs). **(C)** Stacked box plot showing the coherence of allelic effects on mRNA and plasma protein levels by shared cis-e/pQTLs. **(D-F)** Boxplots showing examples of shared cis-e/pQTLs with positively coherent (D, FOLR3), non-coherent (E, IL6R) and negatively coherent (F, MICA) allelic effects on tissue transcript and plasma protein levels. **(G)** Scatterplot comparing the maximal and minimal beta values of the allelic effects of eQTLs across tissues compared to corresponding plasma pQTLs. If the maximum beta value is negative, both eQTL and pQTL have a negative effect and are negatively coherent. If the maximum beta value is positive and greater than the absolute difference between the betas, both eQTL and pQTL have a positive effect and are positively coherent. If the maximum beta value is positive and smaller than the absolute difference between the betas, then the smallest beta must be negative and the eQTL and pQTL have divergent effects. **(H)** UpSet plot showing number of plasma cis-pQTLs colocalizing with cis-eQTLs across tissues. **(I)** Venn diagram showing the number of plasma cis-pQTLs colocalizing with tissue cis-eQTLs and GWAS lead SNPs for cardiometabolic traits. **(J)** Dot plot showing distribution of colocalizing tissue eQTLs and plasma pQTLs across the genome (PP.H4 > 0.5). Color coded dots are also co-localized with lead SNPs identified by GWAS for CAD, LDL, BMI and CRP. Dot size reflects the posterior probability of a shared causal SNP.

In contrast, colocalization analysis (PP > 0.5) revealed that 51/146 eQTL/pQTL pairs were tissue-specific (Figure 2F) (Supplementary Table 3) with liver-specific pairs standing out including 27 unique plasma proteins (AMN, CASA5, CCL16, CD8A, CDCP1, CDHR5, CFHR5, CPE, DDR1, DKK3, ENPP7, F11, F7, FCN2, IL17RB, IL1RL2, IL4R, LILRA5, NAAA, NPRXR, OSMR, PAMR1, PVRL2, SAA4, SULT2A1, THBS2 and TIMD4). Of the 144 unique plasma proteins with colocalizing eQTLs/pQTLs in at least one tissue (Figure 2F), eight pQTLs also colocalized with risk SNPs identified by GWAS for cardiometabolic traits^12–15^ (Figure 2G) (Supplementary Table 4). Whereas the pQTLs for plasma levels of CD40, SMAD5 and SORCS2 (GWAS trait: plasma LDL levels), B3GNT1 (plasma levels of C-reactive protein (h-CRP levels)) and FES (CAD) colocalized with eQTLs acting across several tissues, the pQTLs for IL4R (h-CRP levels) and SEMA3F (body-mass index (BMI)) only colocalized with eQTLs specific to the liver and visceral abdominal fat, respectively (Figure 2G).

### The transcriptional activity of liver “seed-genes” is of particular importance for their plasma protein levels

In STARNET, genetic variation explained 9.4% of the plasma protein levels, which was slightly below recent estimates of 20.5% in 54k individuals of the UK biobank^2^. The remaining ∼80 % portion of the variation in plasma protein levels may at least in part be explained by non-genetic determinants (e.g., micro-environments) of mRNA levels of their corresponding coding genes across tissues (Figure 2). To examine this, we calculated pairwise Spearman correlations between plasma protein levels and corresponding mRNA levels of coding genes across the seven STARNET tissues (Figure 3A). Of 863 plasma proteins that had measurable mRNA levels in at least one STARNET tissue, 446 significant seed gene correlations were identified for 262 unique plasma proteins (FDR 5 %, Figure 3B, Supplementary Table 5). Among these, 101 unique liver seed genes were correlated with a total of 161 plasma proteins. Besides the liver, tissue-specific seed gene-plasma protein correlations were found in blood (n=28), subcutaneous fat (n=21), skeletal muscle (n=13) and visceral abdominal fat (n=11) (Figure 3B, Supplementary Table 5). For seed genes regulated specifically in the arterial wall (AOR and MAM) or by several tissues in parallel, the numbers of correlating plasma proteins per category were <10 (Figure 3B, Supplementary Table 5). Among the 101 unique liver-specific seed gene-plasma protein correlations,18 coincided with the 27 proteins with colocalizing eQTLs/pQTLs (Figure 2F). Given post-translational modification mediated by kinases, phosphatases, transferases and ligases, it is plausible that mRNA of genes other than the seed genes may correlate with plasma protein levels. However, across the STARNET tissues, we found that plasma proteins predominantly correlate with their seed genes rather than with unspecific genes (Figure 3C). Moreover, seed gene-plasma protein correlations were largely concordant in direction (Figure 3D) although there were some examples of negative correlations (Figure 3E). Top-ranked seed genes positively correlating with the levels of their corresponding plasma proteins (Figure 3F) were in order of magnitude; liver *XPNPEP2, AMN, ENPP7, PCOLCE, CCL16, CDHR5, CCL24, GPC5* and *CLSTN2*; skeletal muscle *DHLK1, CHRDL2* and *ADAM23*; aortic arterial wall *FKBP4*; internal mammary artery *TNFRS10B*; visceral fat *OSCAR* and *SCARB2*, subcutaneous fat *FCRL6*, *PROK1*, *CHIT1* and *CD38* and blood *FOLR3*, *LAIR2*, *TCL1A*, *OSM* and *SIGLEC1*. Examples of negatively correlated genes were *CTSH* (liver), *ABL1* (atherosclerotic arterial wall), *IL6R* and *RSPO1* (early atherosclerotic arterial wall), *ARS2* and *FGF2* (visceral abdominal fat) and *LILRB2* (blood) (Figure 3F). Using a subset of STARNET subjects (n=145) with gene expression data available across all seven tissues, we estimated the association between plasma protein levels and tissue-specific seed gene expression after adjusting for the expression of the coding gene in all other tissues in a multivariate model. In this analysis, 38 liver, 11 blood, 6 each of the early atherosclerotic arterial wall, subcutaneous and visceral fat, 4 skeletal muscle, and 2 late atherosclerotic arterial wall seed genes (n=73) remained seed genes significantly correlated with their corresponding plasma proteins (Figure 3G, Supplementary Table 6)

**Figure 3:**
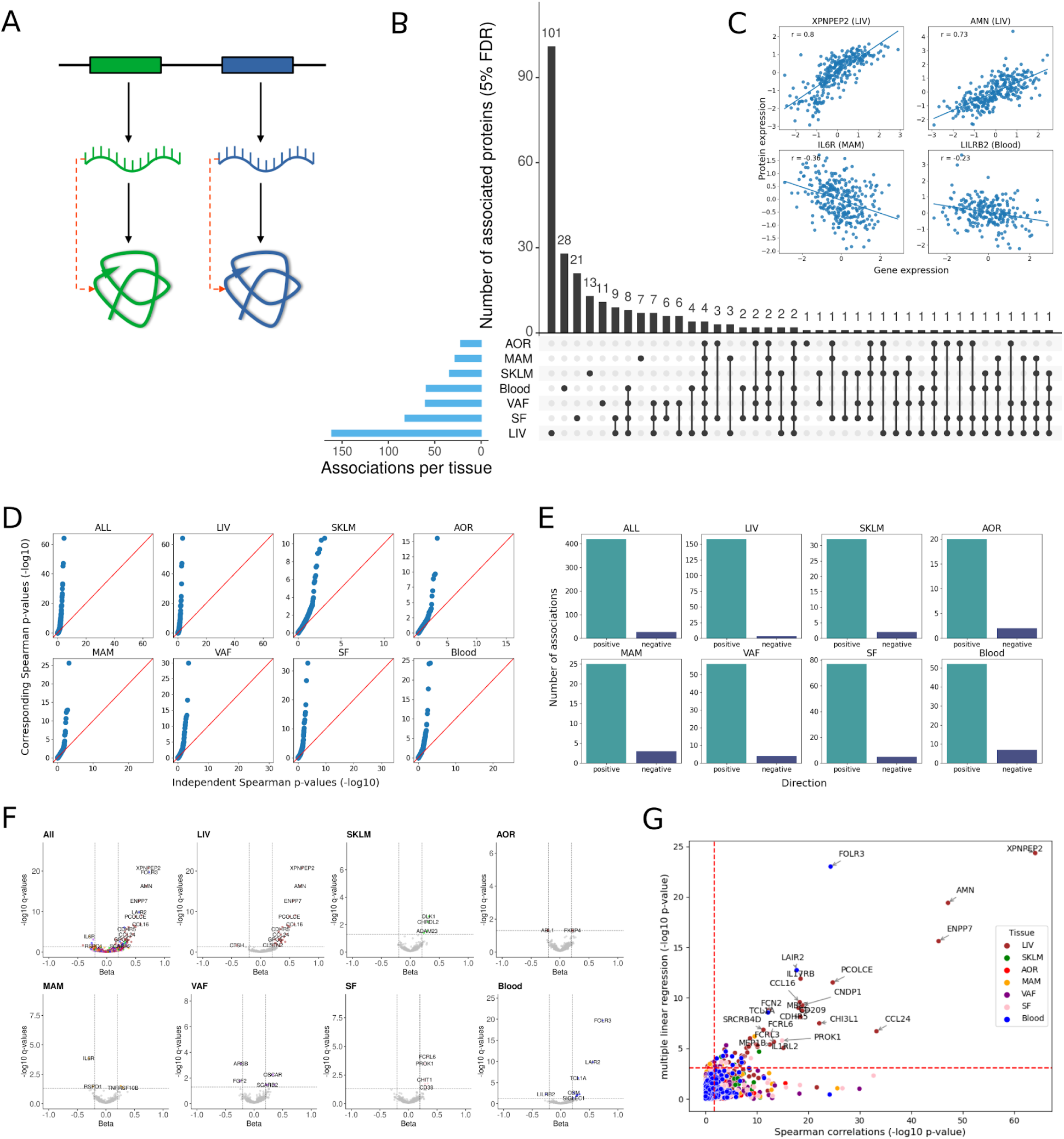
Associations of the expression of seed genes in the arterial wall and metabolic tissues with the levels of their corresponding proteins in plasma. **(A)** Schematic illustration showing the principal hypothesis that the overall (genetic and micro-environmental) regulation of mRNA levels in specific tissues are transmitted to also regulate the levels of corresponding proteins in plasma. Black arrows, flow of information; dashed red arrows, measured correlations. **(B)** Upset plot showing the numbers of tissue-specific transcripts that correlate with their corresponding plasma proteins according to Spearman correlation analysis (q<0.05). **(C)** Scatterplots showing examples of positive (top) and negative (bottom) correlations between levels of tissue-specific seed gene mRNAs and corresponding plasma proteins. **(D)** Quantile-Quantile (Q-Q) plots showing the distribution of p-values for pairwise Spearman correlations between the expression levels of seed genes versus non-seed genes and plasma protein levels across (ALL), and within specific tissues. **E)** Staple grafts showing the number of positively and negatively correlated seed genes with plasma proteins across (ALL), and within specific tissues. **(F)** Volcano plots of Spearman correlations between seed gene mRNA levels across tissues with corresponding plasma proteins. Dashed lines indicate significance thresholds for q-value (q < 0.05) and beta (estimate < -0.2 or > 0.2). Indicated protein IDs (occurring only once) show the tissue-belonging of the strongest seed gene/plasma protein Spearman correlations. **(G)** Scatterplot showing the relationship between tissue-specific Spearman seed gene/plasma protein correlations and seed gene/plasma protein associations in the same tissue adjusted for seed gene expression in all other tissues using multiple linear regression. Red dashed lines indicate 5% FDR thresholds.

### Plasma proteins are integral parts of gene-regulatory networks acting across the arterial wall and metabolic organs

Although the transcriptional activity of some seed genes, particularly in the liver, could be directly linked to their plasma protein levels, the levels of most circulating proteins likely result from more complex processing, which besides synthesis and secretion also involves post-translational modifications and varying rates of clearance. Hence, besides actual seed genes, the combined effects of several other genes may also be involved in regulating the levels of specific plasma proteins. Recently, we analyzed RNAseq data from metabolic tissues and the arterial wall of the STARNET cohort to introduce a mechanistic framework of 224 (135 tissue-specific and 93 cross-tissue) gene-regulatory networks (GRNs)^11^. As part of this work, we implicated secretory factors (among which many are plasma proteins) as part of the explanation why some gene nodes in these GRNs were linked across tissue borders. Now with newly generated plasma protein data from the same cohort, we could evaluate to what extent the 224 GRNs may be involved in the regulation of plasma protein levels (Figure 4A). Using Spearman correlation analysis, we first examined whether the overall activity of each GRN (reflected in their eigengene values (Methods)) were related to the levels of plasma proteins. Out of the 974 measured plasma proteins, remarkably 732 and 727, respectively, were significantly (FDR=5%) associated with at least one of 108 tissue-specific GRNs and 75 cross-tissue GRNs identified in STARNET (Figure 4B, Supplementary Table 7). Thus, only 96 proteins (<10% of the measured proteins) lacked associations with seed genes or GRNs (Figure 4C). As judged from the strongest Spearman Rank associations between the levels of plasma proteins and seed genes or GRN activity, cross-tissue GRNs had the most associated plasma proteins (n=369), followed by blood-specific GRNs (n=279), isolated seed genes (n=103) and liver-specific GRNs (n=68) (Figure 4C). If only considering seed gene-associated plasma proteins (n=382, FDR<5%), 191 plasma proteins were uniquely associated with at least one seed gene in one tissue not belonging to a GRN, 60 uniquely with seed genes in cross-tissue GRNs and 44 uniquely with seed genes in tissue-specific GRNs (Figure 4D). 51 plasma proteins were both associated with the seed gene and with the cross-tissue GRN it belonged to. For tissue-specific GRNs, the corresponding number was 26. 22 plasma proteins were associated with both cross-tissue, and tissue-specific GRNs (Figure 4D).

**Figure 4:**
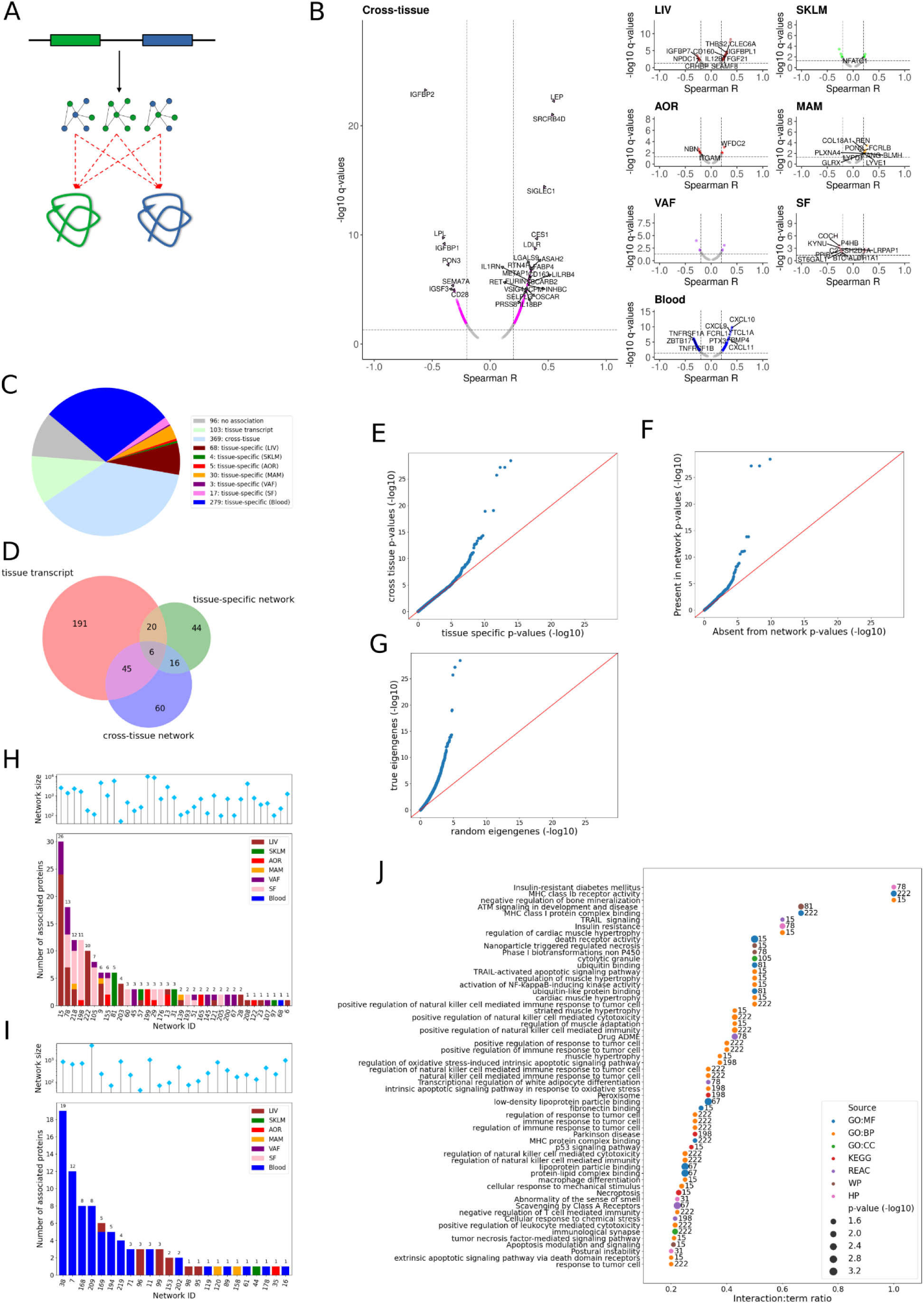
Associations of gene-regulatory networks in the arterial wall and metabolic tissues with levels of plasma proteins. **(A)** Schematic illustration showing the principal hypothesis that the overall activity of gene-regulatory networks (i.e., eigengene values) acting within (i.e., tissue-specific) or across (i.e., cross-tissue) the arterial wall and metabolic tissues are regulating the levels of plasma proteins. Black arrows, flow of information; dashed red arrows, measured correlations. **(B)** Volcano plot showing the pairwise correlations between the eigengene values of 224 GRNs in STARNET^11^ and 974 proteins measured in plasma. Dashed lines indicate significant thresholds for Spearman correlation significance (q < 0.05) and coefficients (Spearman R < -0.2 or > 0.2). Colors indicate whether the indicated GRNs are tissue-specific (>95% of GRN nodes from one tissue) or cross-tissue (<95% of the GRN nodes from one tissue). Indicated protein IDs (only occurring once) show the tissue-belonging of GRNs (i.e., cross-tissue or tissue-specific) having the strongest correlation to a given plasma protein. **(C)** Pie Chart showing fractions of the top-ranked Spearman Rank correlations for each of the measured 945 plasma proteins with either tissue-specific seed genes, tissue-specific GRNs, or cross-tissue GRNs **(D)** Venn diagram showing the overlap of plasma proteins with Spearman Rank associations (FDR 5%) with seed genes, tissue-specific GRNs and cross-tissue GRNs containing the seed genes . **(E)** Q-Q plot showing the distribution of p-values for cross-tissue, vs. tissue-specific GRN/plasma protein Spearman rank correlations. **(F)** Q-Q plot showing the distribution of p-values for GRN/plasma protein Spearman rank correlations fort cross-tissue GRNs containing vs. not containing the corresponding seed gene. **(G)** Q-Q plot showing the distribution of p-values for associations with true vs permuted eigengenes. **(H)** Stacked histogram showing the number of plasma proteins associated with individual cross-tissue GRNs. The tissue-belonging of the seed gene(s) in the networks corresponding to the associated plasma protein(s) are color-labeled. The overarching lollipop chart shows the number of gene nodes in each GRN. **(I)** Stacked histogram showing the number of plasma proteins associated with individual tissue-specific GRNs. The tissue-belonging of the seed gene(s) in the networks corresponding to the associated plasma protein(s) are color-labeled. The overarching lollipop chart shows the number of gene nodes in each GRN. **(J)** Horizontal bar plot showing the 30 top-ranked GO or pathway annotations for the plasma proteins regulated by specific cross-tissue GRNs. GRN IDs^11^ are indicated at the end of each bar. GO, gene ontology; BP, biological process; MF, molecular function; CC, cellular compartment; KEGG, Kyoto Encyclopedia of Genes and Genomes Pathways; REAC, Reactome pathway database; WP, Wikipathways and HP, Human phenotype ontology.

Interestingly, we found that the plasma protein associations with the cross-tissue GRNs were systematically much stronger than the associations with the tissue-specific GRNs (Figure 4E). In fact, of the 732 plasma proteins associated with one or more of the 108 tissue-specific GRNs, 404 had a stronger association with at least one cross-tissue GRN. The strongest associations with cross-tissue GRNs included the plasma proteins CES1, LDLR, LEP, LPL, IGFB1, IGFB2, SIGLEC1 and SRCRB4D (Figure 4B). Additionally, we found that the cross-tissue GRN-plasma protein associations were stronger when the seed genes were present in the GRN compared to when they were absent (Figure 4F). There were 36 cross-tissue GRNs in which the seed genes to the associated plasma proteins were present in the network (Figure 4G, upper panel). Nine of these cross-tissue GRNs were associated with > 5 plasma proteins (Figure 4G, upper panel). The tissue-belongings of seed genes present in these cross-tissue GRNs varied but were mainly found in the liver followed by subcutaneous and visceral fat (Supp Table 7). Some seed genes in these cross-tissue GRNs were active in more than one tissue, predominately in combinations of visceral and subcutaneous fat, and liver (Supp Table 7). In addition to the cross-tissue GRNs, there were 24 tissue-specific GRNs where the seed genes to the associated plasma proteins were present in the network. Of these, the seven GRNs associated with >4 plasma proteins were all blood-specific (Figure 4G, lower panel). A majority of these blood-specific GRNs as well as their associated plasma proteins involved regulation of immune responses (GO:0050776) with strong clinical associations to the number of diseased vessels in the CAD patients.

### Plasma protein-associated cross-tissue GRNs are critical for cardiometabolic diseases

In terms of the number of plasma proteins and their correlation strengths with GRNs, the cross-tissue GRNs stood out (Figure 4B). Specifically, we identified 9 cross-tissue GRNs for which there were > 5 associated plasma proteins/GRN (Supplementary Table 8) in which the corresponding seed genes also were present (Figure 4G). The biological processes and pathways associated with these cross-tissue GRNs showed high relevance for several cardiometabolic disorders like obesity/insulin resistant/diabetes mellitus (GRN78), cardiac muscle hypertrophy (GRN15), leukocyte activation (GRN222/GRN105), apoptotic signaling/oxidative stress (GRN198/GRN155), immune response/inflammation (GRN218), cellular lipid processing (GRN81) and LDL particle binding (GRN67, <u>starnet.mssm.edu/module/67</u>).

Two of these cross-tissue GRNs (GRN105/GRN222) were smaller in size (n= 111/178) and had near tissue-specific properties; GRN105 with 91% of its nodes in the subcutaneous fat (<u>starnet.mssm.edu/module/105</u>) and GRN222, (n=178) with 89% of its nodes in the liver (<u>starnet.mssm.edu/module/222</u>). Both these GRNs involved genes regulating *leukocyte activation* (GO:0045321, *P*=1.1180e-57 (GRN105) and *P*=8.8533e-81 (GRN222)), which also included their associated plasma proteins; GRN105-specific GNLY, GZMH, GRN222-specific CRTAM, SLAMF, CD48, LY9, CD160; and those common to both these GRNs; KLRD1, FCRL6, CD6, CD5, GZMA (Supplementary Table 8). Interestingly, besides GRN105 and GRN222 controlling leukocyte activation in the subcutaneous fat and liver, respectively, there were highly similar GRNs controlling leukocyte activating in other tissues, including GRN174 in the early atherosclerotic arterial wall (n=439, GO:0045321; *P*=1.3739e-136, <u>starnet.mssm.edu/module/174</u>), GRN122 in the advanced atherosclerotic arterial wall (n=766, GO:0045321; *P*=6.7588e-140, <u>starnet.mssm.edu/module/122</u>), GRN 168 in blood (n=710, GO:0045321; *P*=3.0503e-52, <u>starnet.mssm.edu/module/168</u>). GRN54 in skeletal muscle (n=119, GO:0045321; *P*=6.2156e-44, <u>starnet.mssm.edu/module/54</u>), and GRN121 primarily in visceral abdominal fat (n=1052, GO:0045321; *P*=2.4403e-114, <u>starnet.mssm.edu/module/121</u>). Notably, these seven GRNs shared several top key drivers like *BCL11B* and *IKZF3* (in all but GRN54) and *IKZF1*, *RUNX3* and *EOMES* (in 5/7 of the GRNs) but also contained rather exclusive key drivers only occurring in 2 of the seven GRNs like *APOBEC3G, GNLY, HLA-DQA1, KLRD1, LEF1, LGALS17A, SCML4, SPIB, TFEC* and *TRAF3IP3*. Thus, there appears to be both tissue-specific, and generic regulation of leucocyte activation across tissues.

Another pair of the nine cross-tissue GRNs regulating > 5 plasma proteins with similar tissue properties and pathophysiological functions were GRN155 (n=1025, <u>starnet.mssm.edu/module/155</u>) and GRN198 (n=624, <u>starnet.mssm.edu/module/198</u>). Both these GRNs were governed by 18-21 subcutaneous fat key drivers but also contained a substantial number of network nodes but no key drivers in the atherosclerotic arterial wall (15-25%). The pathophysiological functions of GRN155 and GRN198 involved *protein processing* like *protein ubiquitination/modification* (GO:0016567, P= 1.1455e-42) and *protein translation* (GO:0006412, P= 4.7222e-66). Each GRN contributed to 4.6% and 6.7% of CAD heritability and both were strongly associated with BMI, waist/hip ratio and plasma levels of hCRP, triglycerides, cholesterols (LDL/HDL) and HbA1c (-log10 *P*=100). The plasma proteins and corresponding seed genes that supposedly mediate *protein processing* signaling from the subcutaneous fat to the atherosclerotic arterial wall by these two GRNs were ADAM23, ACP6 BCL2L11, CD40, CXCL5, DCTN2, FKBP1B, HTRA2, LAMA4, LGALS3, MANF, PARP1, PRDX1, PVALB, QDPR, SERPINB6, SOD1 and TRIM21 (GO:0006979: *cellular response to oxidative stress*). *NFAT5* was identified as a top key driver in both GRN155 and GRN198.

Yet another pair of these nine cross-tissue GRNs with similar tissue proportion and pathophysiological functions were GRN218 (n=2342, <u>starnet.mssm.edu/module/218</u>) and GRN15 (n=2599, <u>starnet.mssm.edu/module/15</u>). While both these networks mainly contained liver nodes (49-90%), GRN218 was governed by only 4 subcutaneous fat key drivers (CPVL, IKZF1, LGI2 and MS4A4A) and GRN15 by 8 visceral abdominal fat key drivers (ATF3, CREM, EGR1, ETS2, FOS, FOSL1, FOSL2 and MYC) together with 4 key drivers in the liver (ELF3, IRF1, MT1DP and MT1E). The functions of these GRNs involved *inflammatory responses* (GO:0006954, P= 5.9118e-79) and *responses to cytokines* (GO:0034097, P= 3.1605e-96), respectively. GRN218 and GRN15 contributed to 14% and 13% of CAD heritability and were both strongly associated with BMI, Waist/Hip ratio, the plasma levels of hCRP, triglycerides, cholesterol (LDL/HDL), HbA1c and the number of coronary lesions (-log10 *P*=100). The plasma proteins and corresponding seed genes (CD163, CRHBP, F7, IL18, KYNU, MSR1, PON3, RET, TNFRSF10A and TNFRSF11A in GRN218 and ADM, ANGPTL4, CASP8, CCL3, CD274, CHI3L1, CLEC5A, CSF1, CTSL, EZR, FGF23, FSTL3, IL4R, METRNL, PARP1, PLAUR, SDC4, TGFA, TNFRSF1A, TNFRSF1B, TNFRSF10A, TNFRSF10B, TNFRSF12A, TNFRSF19, TNFSF14 and VEGFA in GRN115, Supplementary Table 8) mediated inflammatory (GRN218) and programmed cell death (GRN115) signaling predominately in the direction from fat to the liver.

### The Adipose-Liver Plasma LeptIN-regulatory network 78 (ALPLIN78) and its top key driver Ubiquitin C-Terminal Hydrolase L1

The cross-tissue GRN with the second highest number (n=13) of associated plasma proteins with seed genes present in the network was ALPLIN78 (Figure 5, n=1403, <u>starnet.mssm.edu/module/78)</u>. The tissue locations of seed genes in cross-tissue GRNs point to the tissues that may be the most important for their subsequent secretion or leakage into plasma and thus, for their plasma levels. For four of ALPLIN78’s associated plasma proteins; CA3 (Carbonic Anhydrase 3), CES1 (Carboxylesterase 1), HSD11B1 (Hydroxysteroid 11-Beta Dehydrogenase 1) and LEP (Leptin), the corresponding seed genes were present in both the subcutaneous and visceral abdominal fat sections of ALPLIN78. In addition, the seed gene to ALPLIN78-associated plasma protein PON3 (Paraoxonase 3) was present in both the subcutaneous fat and liver sections of ALPLIN78. The seed genes of the additional eight ALPLIN78-associated plasma proteins were only observed in one tissue; (*CRHBP*, *Corticotropin Releasing Hormone Binding* Protein/subcutaneous fat; *FCN2*, *Ficolin 2* /visceral abdominal fat and *CNDP1, Carnosine Dipeptidase 1*; *IGFBP2, Insulin Like Growth Factor Binding Protein 2; IL17RB, Interleukin 17 Receptor B; INHBC, Inhibin Subunit Beta C; MEP1B; Meprin A Subunit Beta and PLIN1; Perilipin 1/liver*).

**Figure 5:**
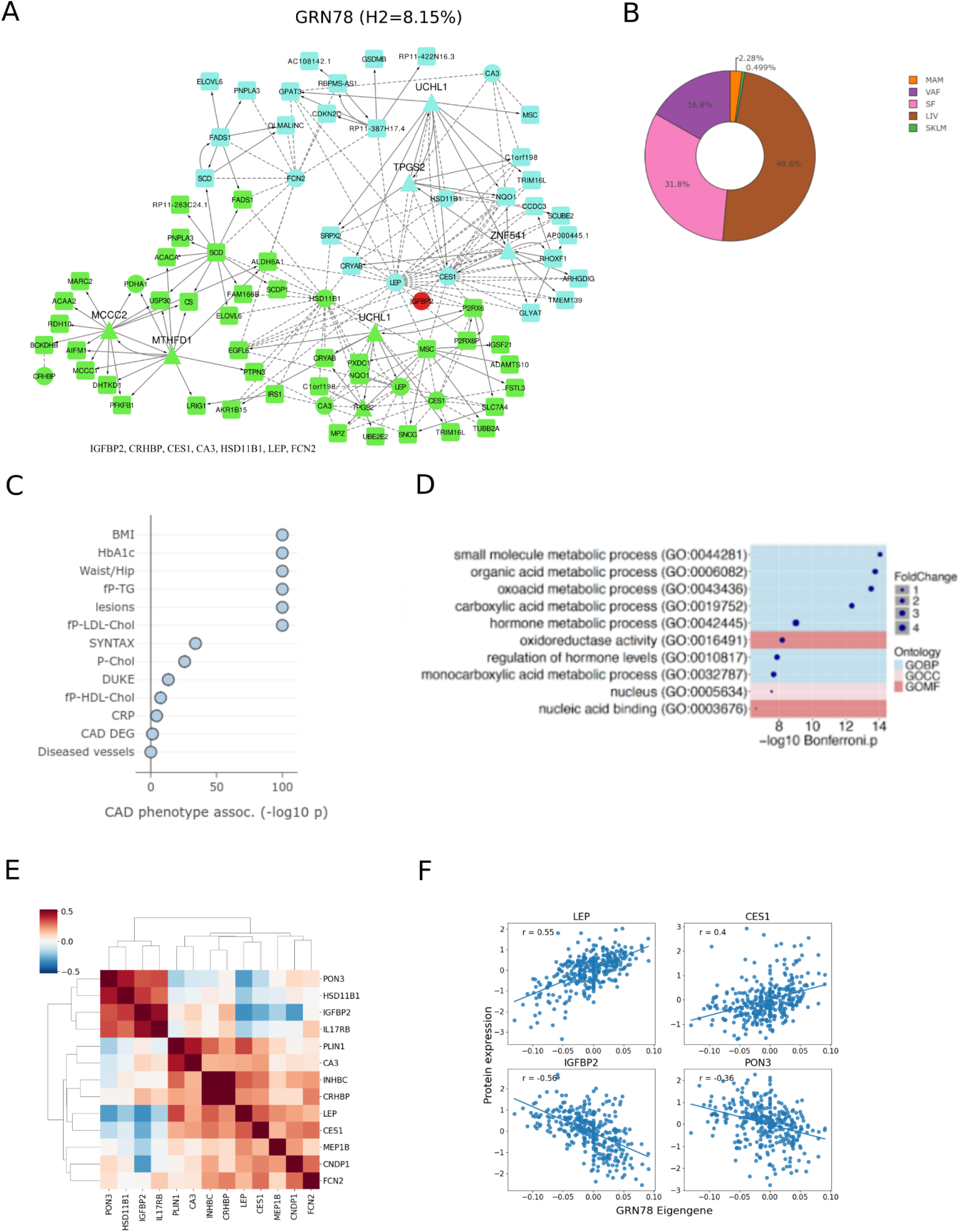
Characteristics of the Adipose-Liver Plasma LeptIN-regulatory network 78 (ALPLIN78) (A) Illustration of ALPLIN78 (i.e., GRN78) showing the first and second neighbors to its 5 key drivers; MCCC2, MTHFD1, TPGS2, UCHL1 and ZNF541 including co-expression edges to nodes corresponding to GRN78-associated plasma proteins^11^. (B) The tissue belonging to ALPLIN78 gene nodes. MAM, early atherosclerotic mammary artery; VAF, visceral abdominal fat; SF, subcutaneous fat; LIV, liver and SKLM, skeletal muscle^11^. (C) Clinical associations of ALPLIN78 with CMD and CAD phenotypes. -log10 P-values represent enrichments of ALPLIN78 with genes associated with indicated phenotypes (Spearman Rank associations, *P*<0.05). BMI, body-mass index; fP, fasting plasma; TG, triglycerides; lesions, number of coronary lesions; Chol., cholesterol; LDL, low-density lipoprotein; HDL, high-density lipoproteins; CRP, high-sensitivity C-reactive protein; CAD DEG, differential expressed genes between CAD-cases and CAD-free controls; Diseased vessels; number of diseased coronary vessels^11^. (D) Top gene ontologies (GO) of ALPLIN78 nodes. GOBP, biological processes; GOMF, molecular functions and GOCC, cellular components^11^. (E) Heatmap of pairwise Spearman correlations between the 13 plasma proteins associated with the ALPLIN78 eigengene (CA3, CES1, *CNDP1,* CRHBP, FCN2, HSD11B1, IGFBP2, IL17RB, INHBC, LEP, MEP1B; PLIN1 and PON3). Proteins have been ordered by hierarchical clustering. (F) Scatter plots between ALPLIN78’s eigengene value and its four most associated plasma proteins according to statistical significance. Spearman correlation coefficient (r) values are indicated in the plot.

ALPLIN78 also stood out for several other reasons including its 8.15% CAD heritability contribution and content of BMI, CAD, LDL and HbA1c GWAS candidate genes (Supplementary Table 9), top-regulatory role of fat genes (Figure 5A), clear cross-tissue nature (Figure 5B), firm correlations to cardiometabolic traits including atherosclerosis severity (Figure 5C) likely mediated through its overarching pathophysiological function (Figure 5D). Despite that near 50 % of ALPLIN78’s nodes were found in the liver, this cross-tissue GRN only had five key drivers, which all were found in either subcutaneous, or visceral abdominal fat (Figure 5C), suggesting a role of ALPLIN78 in largely mediating signaling from adipose to govern gene activity in the liver. In addition out of these six key drivers (UCHL1(VAF/SF), ZNF541 (VAF), MTHFD1 (SF), TPGS2 (VAF), MCCC2 (SF)), the top key driver *Ubiquitin C-Terminal Hydrolase L1* (*UCHL1*) (FDR=1.2326e-48) was found in visceral abdominal fat but interestingly appeared again as ALPLIN78’s 5^th^ highest ranked key driver; UCHL1 in subcutaneous fat (FDR=5.9688e-29). The correlations of individual plasma proteins with the ALPLIN78 eigengene value indicated that some of the 13 plasma proteins were co-regulated whereas others were not (Figure 5E). Among the four most associated plasma proteins, LEPT and CSN1 were positively, and IGFBP2 and PON3 negatively, correlated with ALPLIN78 (Figure 5F).

## DISCUSSION

By screening 974 plasma proteins in 532 well-characterized, multi-tissue and data-rich STARNET individuals with cardiometabolic disease^11^, we positioned ourselves to study the influence of genetics, seed genes and GRNs in major metabolic organs and the arterial wall at the levels of plasma proteins. Of 1,860,901 SNPs located in the ±500Kb cis-window around the transcription start sites of 945 genes corresponding to the 974 measured plasma proteins, 50,106 SNPs were identified as cis-pQTLs for 708 unique plasma proteins (FDR=5%). 220 of these proteins had an overlapping cis-eQTL in at least one tissue of which 136 colocalized. By assessing the expression levels of 6,846 seed genes (7 tissue x 974 genes), 262 plasma proteins were found to be associated with their corresponding seed genes in at least one tissue (FDR=5%), whereof 101 were unique to the liver. Most striking, however, was the observation that out of 224 GRNs previously established in STARNET^11^, as many as 183 had at least one significant association with 851 of the 974 proteins measured in plasma (FDR 5%). The highly significant associations between the activity of these GRNs, and the levels of plasma proteins effectively establishes a framework of endocrine signaling integrating metabolic and inflammatory gene activity across major metabolic organs and the arterial wall driving CMDs and CAD. Among these GRNs, the cross-tissue GRN ALPLIN78 stood out and should serve as a future target to prevent obesity and type 2 diabetes possibly by targeting *UCHL1* in fat.

Eight genes with colocalizing tissue cis-eQTLs and plasma cis-pQTLs also colocalized with established risk SNPs for CMDs and CAD. Most notably, for the GWAS trait of plasma LDL, four candidate genes were identified: First, the genetic regulation of CD40 levels in plasma and its gene expression across all tissue but skeletal muscle colocalized with the inherited risk of increased plasma LDL levels. CD40 is a critical macrophage surface receptor protein involved in immune responses, that also plays critical roles in both fatty acid oxidation and glutamine metabolism^16^. Secondly, the cis-pQTLs for the signal transduction protein SMAD5, known for its involvement in hematopoiesis and osteogenesis^17^, were found to colocalize with plasma LDL and cis-eQTLs in both visceral and subcutaneous fat as well as in liver and blood. Thirdly, the plasma cis-pQTLs for SORCS2, a member of the VPS10 domain receptor family involved in intracellular trafficking and signalling^18^, colocalized with LDL levels and cis-eQTLs in in both visceral and subcutaneous fat as well as the liver. Last, DDK3, a secreted glycoprotein that has been demonstrated to reduce cardiac hypertrophy and improve cardiac function in mouse models^19^, colocalized with LDL and cis-eQTLs specifically in the liver.

In addition to LDL levels, the inherited risk of increased plasma CRP levels colocalized with the plasma pQTLs of B3GNT1, a glycosyltransferase implicated in Walker–Warburg syndrome^20^, and its corresponding eQTLs across all tissues but blood. In addition, the genetic regulation of plasma levels of IL4R and CRP colocalized together with its cis-eQTLs specifically in the liver. IL4R is well known for its role in the IL-4 signaling pathway and has been implicated in both tissue inflammation and asthma^21^. In addition for BMI, the plasma levels of SEMA3F implicated in vascular regulation as an inhibitor of angiogenesis^22^, was found to colocalize with both the risk of increased BMI and *SEMA3F* cis-eQTLs specifically in visceral abdominal fat. Finally for CAD, the plasma pQTLs of FES, known for its role to protect against atherosclerosis^23^, was found to colocalize with its eQTLs in liver, visceral abdominal fat and the atherosclerotic arterial wall.

Besides the genetic effects of eQTLs, mRNA levels are affected by tissue-specific micro-environments. Thus, the expression of coding genes across tissues may effectively act as seed genes for their corresponding plasma proteins. We found that coding genes in the liver were more likely to be seed genes followed by coding genes in circulating blood cells. A prominent liver seed gene example was XPNPEP2 (X-Prolyl Aminopeptidase 2, Figure 3C), a membrane-bound metalloprotease which catalyzes the removal of a penultimate prolyl residue from the N-termini of peptides, such as Arg-Pro-Pro, a process suggested to play a role in the metabolism of the vasodilator bradykinin^24^. Another liver seed gene with strong and independent association to its plasma protein was ADM (Amnion Associated Transmembrane Protein) (Figure 3C). Together with CUBN, ADM forms an endocytic receptor required for efficient absorption of vitamin B_12_^25^. In the early atherosclerotic arterial wall (the mammary artery), the seed gene of the plasma protein IL6R (interleukin 6 receptor) was found negatively correlated with its plasma protein levels (Figure 3C). This is potentially interesting since IL6R is believed to be a key driver as part of the NLRP3 inflammasome driving atherosclerosis^26^.Taken together, our observation of seed genes exemplified by the plasma levels of XPNPEP2, ADM and ILR6 that closely mirror their levels of gene expression may have potential as biomarkers for clinically relevant conditions such as in the three examples: Vasodilation (XPNPEP2), Vitamin B_12_ deficiency (ADM) and degree, or resilience to atherosclerosis (ILR6).

In 2022, we published a mechanistic framework for CAD and CMDs that consisted of 224 GRNs acting across major metabolic organs and the arterial wall explaining 60% of CAD heritability beyond the 20% CAD heritability already identified by GWAS^11^. In addition, the statistical significance of CMD and CAD trait associations of these GRNs markedly exceed that of individual genes. In this study, initial analyses indicated that endocrine signaling was likely to be a major underlying cause of this, and particularly for cross-tissue GRNs this explained why some networks have gene-gene interactions acting across tissue borders. Apart from a handful endocrine candidates experimentally validated in mice^11^, for the most part we could only speculate about the underlying signaling system that explained the 89 cross-tissue GRNs identified in STARNET. In the current study, we extended the STARNET tissue data repository with parallel plasma proteomic data from mostly the same individuals and found that 851 of the 974 proteins measured in plasma were associated at least once with the activity of 183 of these 224 GRNs including 75 of the 89 cross-tissue networks displaying the strongest associations with plasma proteins. Rigorous permutation testing ruled out that this could have happened by chance (Figure 4). The remarkable number of highly significant GRN eigengene-to-protein associations compared to that of individual gene-to-protein associations, also has support in the so-called “wisdom of the crowd” effect^27^. This theory states that for true crowds (e.g., demonstrators rather than randomly gathered groups, like people in a subway), the behavior of each individual (e.g., the expression of individual genes) is a weak predictor of down-stream events (e.g., the levels of plasma proteins). In contrast, if a crowd can be accurately identified like the STARNET GRNs, it becomes a strong predictor of downstream events. Indeed, we have previously shown^27^ that across a wide range of independent states, the best consensus predictor is often the first principal component of the individual predictors, i.e. the module eigengene. Thus, overall, it appears plasma proteins are regulated as integral parts of mainly cross-tissue GRNs acting within and across major metabolic organs and the arterial wall.

The concept of GRNs fills two critical knowledge gaps: i) the knowledge gap of molecular interactions combining the general functions of organ systems (e.g. the liver) and their individual pathways (e.g., cholesterol synthesis) mostly reflected in tissue-specific GRNs, and ii) the knowledge gap of molecular interactions taking place in-between organs systems (e.g., adipose-liver) reflected by the cross-tissue GRNs. By learning how the connectivity of these GRNs changes alongside aging and complex disease development, thereby turning organ physiology into pathology, we can effectively start implementing the future of clinical medicine; *clinical systems medicine.* In such future, we will no longer treat CMDs, like obesity, type 2 diabetes and CAD, blindly by targeting isolated genes with little or no follow up, but instead modify the activity of entire GRNs like ALPLIN78 by targeting GRN-specific key drivers, like *UCHL1,* and then monitoring the results of such interventions by measuring changes to GRN biomarkers in blood, like in the case of ALPLIN78, monitoring the combined changes to the plasma levels of CA3, CES1, CNDP1, CRHBP, FCN2, HSD11B1, IGFBP2; IL17RB, INHBC, LEP, MEP1B, PLIN1 and PON3.

## Data Availability

Access request for STARNET is possible through dbGaP (accession number phs001203.v4.p1). Requests for further information and resources should be directed to and will be fulfilled by the lead contact, J.L.M.B.

## RESOURCE AVAILABILITY

All code used in this study is available at the following repository https://github.com/sbankier/STARNET_plasma_proteomics. Access request for STARNET is possible through dbGaP (accession number phs001203.v4.p1). Requests for further information and resources should be directed to and will be fulfilled by the lead contact, J.L.M.B.

## ACKNOWLEDGMENTS

J.L.M.B acknowledges support from the Swedish Research Council (2018-02529 and <u>2022-00734</u>), the Swedish Heart Lung Foundation (2017-0265 and 2020-0207), the Leducq Foundation AtheroGen (22CVD04) and PlaqOmics (18CVD02) consortia; the National Institute of Health-National Heart Lung Blood Institute (NIH/NHLBI, R01HL164577; R01HL148167; R01HL148239, R01HL166428, and R01HL168174), American Heart Association Transformational Project Award 19TPA34910021, and from the CMD AMP fNIH program. T.M. acknowledges support from the Research Council of Norway (project number 312045), the European Union’s Horizon Europe (European Innovation Council) programme (grant agreement number 101115381), and the L. Meltzers Høyskolefond. J.C.K. acknowledges research support from the NIH (R01HL148167), New South Wales health grant RG194194, the Bourne Foundation, Snow Medical and Agilent.

## AUTHOR CONTRIBUTIONS

S.B, T.M, and J.L.M.B contributed to the conception and design of this research. S.B conducted all formal analyses and visualizations and wrote. M.K conducted additional analyses. J.C.K. assisted with the interpretation of results. H.T. assisted with visualizations. The article was written by J.L.M.B, S.B and

T.M. All authors reviewed the article and approved the submitted version.

## METHODS

### The STARNET study

STARNET includes near to 1500 patients with obstructive CAD who underwent coronary artery bypass grafting (CABG) surgery^10,11^. Whole blood samples were obtained the day prior to surgery, whereas the seven tissue samples were obtained during surgery from the atherosclerotic aortic root (AOR), the non- or early-atherosclerotic internal mammary artery (MAM), liver (LIV), skeletal muscle (SKLM) and visceral abdominal (VAF) and subcutaneous fat (SF). All patients gave written informed consent (ethical approvals: Tartu, Dnr 154/7 and 188/M-12, Mount Sinai, IRB-20-03781). Patients with other severe systemic disease (e.g., active inflammatory disease or cancer) were excluded. The extent of coronary atherosclerosis (e.g., SYNTAX score (68, 69)) was assessed from preoperative angiograms. Each participant completed a questionnaire to assess disease history, current drug regimens, and lifestyle (e.g., daily activity, alcohol consumption, smoking) and was screened for 114 clinical variables including serum biochemistry. 31% of the STARNET individuals are females; 32% have diabetes, 75% have hypertension, 67% have hyperlipidemia; and among the cases, 33% had a myocardial infarction before age 60.

### Genotyping and imputation

STARNET genotype data underwent Quality Control (QC) and normalization as described previously^10^. The Human OmniExpressExome-8v1 bead chip was used with GRCh37 and contains 951,117 genomic markers and imputed to 12,450,918 autosomal SNPs. Genotype calls were presented as matrices within the -012 format and a filtering step was included to remove any SNPs which had missing values for any samples and to exclude any SNPs which had a Minor Allele Frequency (MAF) < 5%, resulting in 8,337,055 SNPs.

### RNA sequencing

All STARNET tissues underwent RNA-sequencing as has been previously described^10^. In brief, the RNA-seq data was generated using the HighSeq2000 platform to a depth of 25–40 million reads, with the Ribo-Zero protocol. RNA samples with less than 1M uniquely mapped reads were excluded, which removed 12 samples with extremely low read counts. The read counts of the samples used in the final analysis were between 15-30 million reads. We labelled the transcripts using Ensembl Biomart (GRCh37) with their gene name, chromosome location, gene start, gene end and the Transcription Start Site (TSS).

### Olink proteomics

EDTA plasma samples from the STARNET patients were used for proteomic profiling using the Olink Target96 platform. In brief, this method uses proximity extension assay (PEA) technology^28^, applying two separate antibodies binding to different epitopes of the target proteins. Once bound to a target protein, oligonucleotides situated at the FC domain of the antibody pair hybridize producing a protein-specific DNA-reporter sequence. Quantitative Polymerase Chain Reaction (qPCR) is used to quantify each sequence generating reporter tags working as proxys for the protein contents known as a Normalised Protein eXpression (NPX) values^2,28^. Two runs of proteomic profiling were carried out on plasma from a total of 535 STARNET cases across 11 Olink panels containing a total of 1012 protein assays representing 979 unique proteins. NPX values were adjusted for principal components related to batch effects of the two runs, age and sex using a linear model (Figure S2).

### GWAS summary statistics

GWAS summary statistics used in this study were downloaded from GWAS Catalog^29^. These are described in detail in Table S1.

### Tissue-specific eQTL and plasma pQTL inference analysis

We carried out cis-pQTL discovery across all unique proteins measured, and cis-eQTL discovery for the corresponding genes. The cis loci of the targeted proteins were defined based on chromosome and base pair position. Each Olink ID is linked to a UniProt ID^30^, which in turn was used to annotate each Olink assay (Release 2023_04) with the corresponding gene name. Information from Ensembl Biomart (GRCh37) was then used to annotate each protein with chromosome and base pair position. We defined cis-SNPs for each protein based on a window ± 500 Kb of the Transcription Start Site (TSS) for the corresponding gene and using SNP information from the Single Nucleotide Polymorphism Database (dbSNP)^31^. These cis-SNPs were then mapped to STARNET genotypes which resulted in 1,860,901 genotypes for 945 proteins. Cis-pQTL discovery was carried out using linear regression as implemented by MatrixEQTL^32^. This was performed using the standard procedure as directed by the program authors, however in addition to the default model coefficients, the beta standard error was also calculated. Multiple testing was corrected for using MatrixEQTL’s built-in False Discovery Rate (FDR) correction at a 5% FDR threshold. This was carried out using protein measurements for pQTL discovery and then subsequently using tissue gene expression measurements for eQTL discovery for corresponding mRNA transcripts. We also estimated the variance explained by each cis-pQTL for their associated protein. From all significant pSNPs (FDR = 5%), we selected independent SNPs by grouping SNPs based on LD (Pearson r > 0.8) and selecting the strongest SNP based on p-value. For each protein, we then fitted a linear model in R (version 4.1.2), where the genotypes of the independent SNPs act as the explanatory variables for the protein expression. We then used the adjusted coefficient of determination (r2) as a measure of the variance explained for the impact of cis-pQTLs for that protein.

### Colocalization analysis of tissue-specific eQTLs, plasma pQTLs and genetic loci identified by GWAS

Colocalization between plasma cis-pQTLs and tissue cis-eQTLs in STARNET was estimated using the Approximate Bayes Factor Colocalization method from the Coloc package^33^ (version 5.2.0) in R. A pre-filtering step was first conducted where only proteins that had at least one matching eSNP and pSNP (FDR < 5%) were taken forward for colocalisation analysis. For each transcript-protein pair, QTL summary statistics were obtained for all SNPs within a 100 Kb window of the TSS. Information provided to Coloc for each set of summary statistics included the beta, beta variance, number of samples, rsID, position and Minor Allele Frequency. This returned the posterior probability that both traits are associated and share a single causal variant. Colocalisation was also carried out using the same approach between cis-pQTLs and GWAS summary statistics.

#### Spearman correlations between tissue-specific mRNA and plasma protein levels

Correlations between the plasma protein and gene expression levels of their corresponding coding genes in each tissue were calculated using Spearman’s ranked correlation coefficients. In each tissue, protein and gene expression datasets were first matched based on their STARNET identity. Correlations were calculated for each transcript-protein pair using the spearmanr function from Scipy Stats^34^ (version 1.7.3) in Python (version 3.9.7), returning both the Spearman’s correlation coefficient and corresponding p-value. Multiple testing correction was conducted through the Storey-Tibshirani method of q-value estimation^35^. We compared correlations between proteins-corresponding transcript and proteins-independent transcripts using the Spearman p-value as Quantile-Quantile (QQ) plots on both a tissue-by-tissue basis and for all tissues combined. The term “seed gene” was used for a coding gene within a given tissue that was significantly correlated to its corresponding plasma protein.

#### Multiple linear regression of tissue-specific mRNA and plasma protein levels

The association between protein and transcript expression was estimated using multiple linear regression using the lm function in R (version 4.1.2). Common samples were identified between all transcript and protein expression datasets and samples that did not have information in all datasets were removed, which returned 145 samples. Additionally, proteins were removed in cases where the transcript was not present in any of the STARNET gene expression datasets, resulting in 864 proteins which were taken forward. A linear model was constructed for each protein (Y), where the transcript for each tissue where the expression was present were fitted as covariates (X) (equation 1).

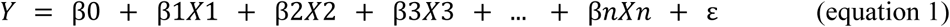

For each protein the coefficients were extracted from the model and multiple testing was corrected for using the Storey-Tibshirani procedure^35^.

### Inference of gene regulatory networks (GRNs)

STARNET tissue gene expression data has been previously used for the inference of co-expression network modules, which have been described extensively in Koplev et al^11^. Networks were constructed using weighted correlation network analysis (WGCNA)^36^, where module inference was conducted within tissues and for all possible pairwise combinations across tissues, for the identification of both tissue-specific and cross-tissue networks. Networks were defined as being tissue-specific based on a purity threshold of 0.95.

### GRN-eigengene Spearman correlations

Using pairwise Spearman correlation, we measured the correlations between the expression levels of all plasma proteins and the eigengenes of all 224 co-expression GRNs, which are principal component measures of the co-expression module gene expression measurements, and were described in Koplev et al^11^. We selected significant correlations based on the Spearman q-value, following multiple testing correction using the Storey-Tibshirani procedure^35^. Spearman correlations were calculated in Python using the spearmanr function from Scipy Stats^34^ (version 1.7.3). We used QQ plots to describe the inflation of p-values between different classes of protein-eigengene correlations i.e. for cross-tissue vs tissue-specific network or for networks containing the corresponding transcript of the correlated protein vs those without. The protein associations were then investigated using the “STARNET browser”, an interactive web portal that provides both functional and structural information about the 224 GRNs, as described in Koplev et al^11^.

## SUPPLEMENTARY MATERIAL

**Figure S1:**
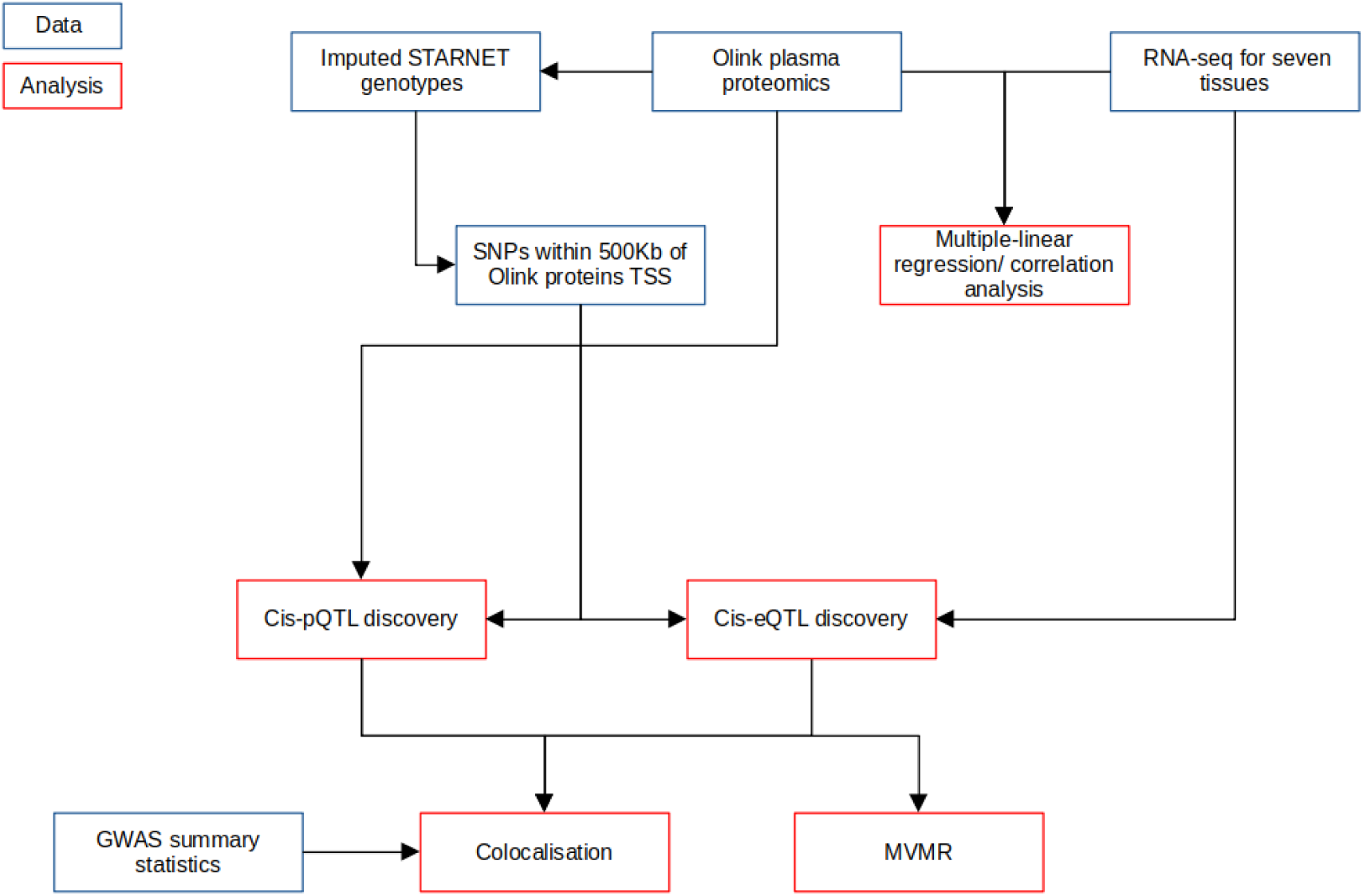
Analysis work-flow diagram. Blue boxes represent data inputs and red boxes indicate analyses. Arrows indicate flow of information.

**Figure S2:**
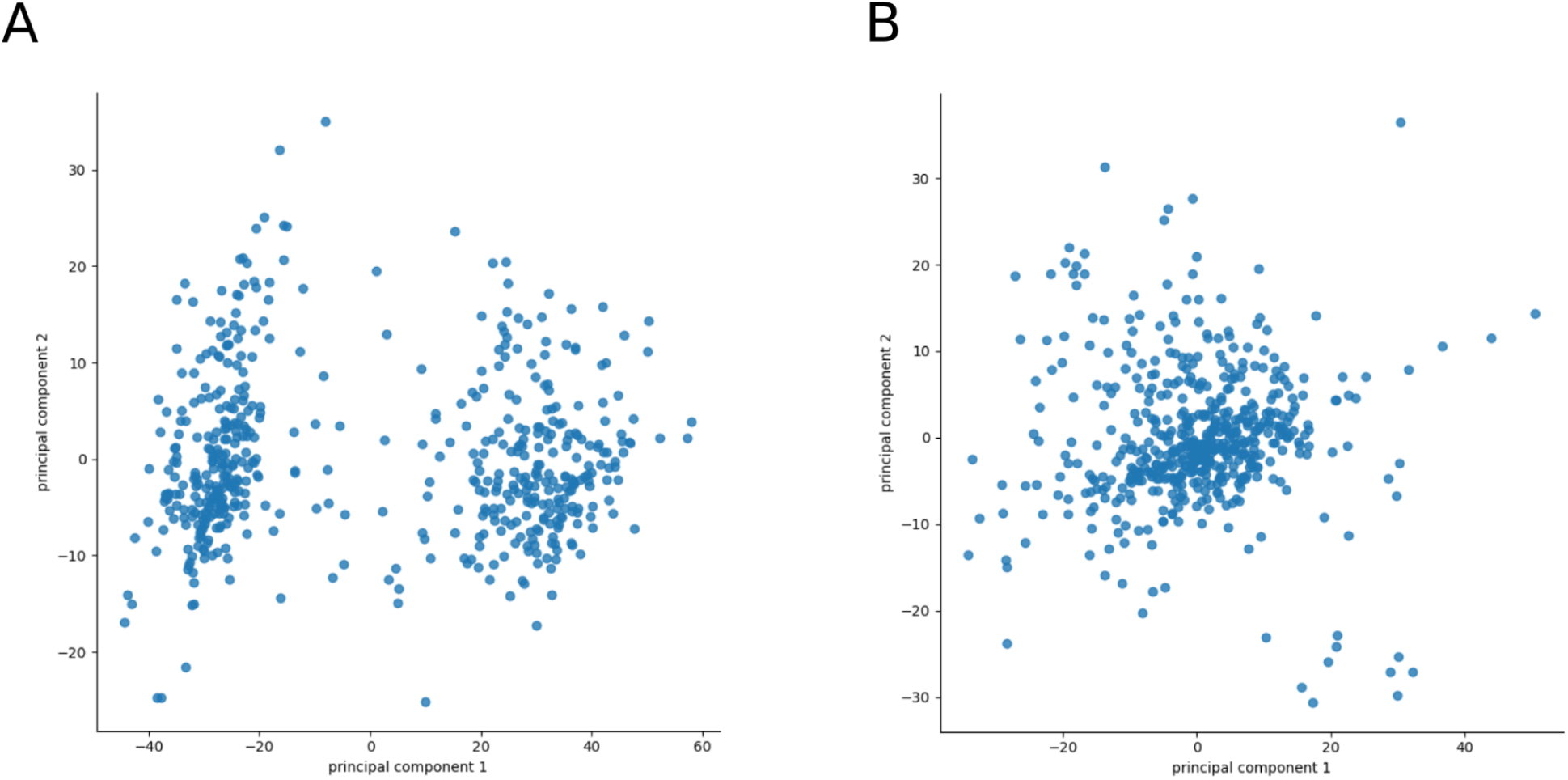
PCA of individual level Olink plasma protein measurements. **(A)** PCA of unadjusted Olink measurements of STARNET samples (n=535). **(B)** PCA of Olink measurements following adjustment for age, sex and the first principal component from the unadjusted data (n=532).

**Figure S3:**
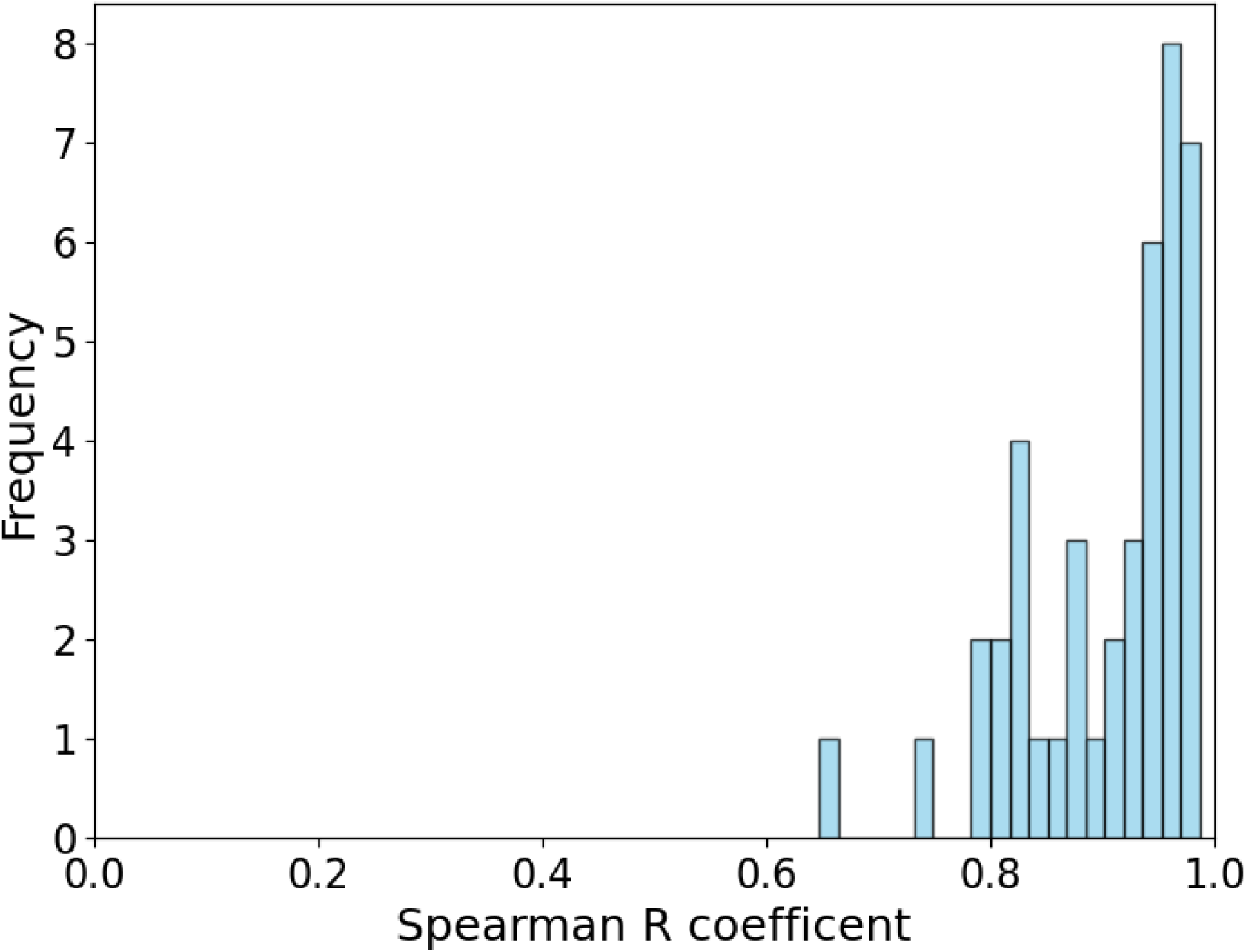
Distribution of pairwise correlations between alternate Olink protein assays for the same protein.

